# Multi-route respiratory infection: when a transmission route may dominate

**DOI:** 10.1101/2020.04.06.20055228

**Authors:** Caroline X. Gao, Yuguo Li, Jianjian Wei, Sue Cotton, Matthew Hamilton, Lei Wang, Benjamin J. Cowling

## Abstract

The exact transmission route of many respiratory infectious diseases remains a subject for debate to date. The relative contribution ratio of each transmission route is largely undetermined, which is affected by environmental conditions, human behavior, the host and the microorganism. In this study, a detailed mathematical model is developed to investigate the relative contributions of different transmission routes to a multi-route transmitted respiratory infection. It is illustrated that all transmission routes can dominate the total transmission risk under different scenarios. Influential parameters considered include dose-response rate of different routes, droplet governing size that determines virus content in droplets, exposure distance, and virus dose transported to the hand of infector. Our multi-route transmission model provides a comprehensive but straightforward method to evaluate the transmission efficiency of different transmission routes of respiratory diseases and provides a basis for predicting the impact of individual level intervention methods such as increasing close-contact distance and wearing protective masks. (Word count: 153)

**Highlights:**

1. A multi-route transmission model is developed by considering evaporation and motion of respiratory droplets with the respiratory jet and consequent exposure of the susceptible.
2. We have illustrated that each transmission route may dominate during the influenza transmission, and the influential factors are revealed.
3. The short-range airborne route and infection caused by direct inhalation of medium droplets are highlighted.

## Introduction

The 2003 Severe Acute Respiratory Syndrome (SARS) epidemics, the 2009 H1N1 influenza (Swine Flu) pandemic, the 2015 Middle East Respiratory Syndrome – coronavirus (MERS-CoV) epidemics and the ongoing novel human coronavirus (SARS-CoV-2) global pandemic have all highlighted the importance of studying the transmission mechanism of respiratory infectious diseases (1–4).

Respiratory diseases are often simply assumed to be transmitted via “close contact”; however, the transmission mechanisms are complex involving more than one transmission route including direct or indirect contact, large droplet, and airborne routes (5–9). There are many physical (respiratory particles and droplets generation), virological (viral loading, survival, location of virus receptor, etc.), behavioral (exposure distance, frequency of handshaking and surface touching, etc.) and environmental factors (temperature, humidity, ventilation, etc.) that affect the transmission (8, 10).

Hence, respiratory infections may show various characteristics under different contact scenarios. For example, airborne transmission was identified as played the leading role in an influenza outbreak on a commercial aircraft in 1977 in Alaska (11).

Conversely, in a H1N1 outbreak in a tour group in China, close contact was the most correlated factor with the transmission (12). Conflicting evidence for transmission routes, like these two cases, are prevalent for almost all respiratory infectious diseases (8). Failure in understanding the complex multi-route transmission mechanisms leads to recommendations of more conservative intervention methods such as keep a distance rather than increasing ventilation and wearing masks. The consequences of more conservative interventions can be catastrophic such as the global pandemic of SARS-CoV-2 outbreak (13–15).

However, understanding of multi-route transmission is by no means an easy task. Findings from animal challenge models are difficult to extrapolate to human transmission (16). Human challenge models are expensive and often unethical (17). Observational studies of existing outbreaks often fail to capture important time relevant evidence. A more feasible approach is to use mathematical models to describe the multi-route transmission using known parameters such as droplet generation rate, virus shedding rate, and virus survival rates. A few mathematical studies have developed multi-route transmission models such as by Nicas and Jones (18), Atkinson and Wein (19) and Spicknall and colleagues (20). However, many critical factors, such as evaporation of respiratory droplets, travelling of the large droplets in the respiratory jet, pulmonary deposition, dose-response rate for different route were not fully evaluated in these models, which may underestimate the role of smaller droplets.

In this paper, we first provide comprehensive definitions of transmission routes which incorporates the underlying physical principles of multi-route transmission. Second, we establish a more advanced simulation model to describe the infection via different routes considering the physical components of particles and droplets movement, differences in possible viral dose-effect as well as human-behavior factors such as touching mouth and nose. We use influenza as an example due to the large number of related studies for extracting modelling parameters. Using the model, we aim to challenge the traditional dichotomous thinking of close contact transmission vs airborne (aerosol) transmission via highlighting the scenarios under which each transmission route may dominate and how environmental and behavior factors interact in the transmission mechanism.

## Methodology

### Transmission routes definitions

Traditional definitions for transmission routes include the airborne route (also referred as aerosol transmission) (21), large droplet route, and contact route (6, 22). However, such definitions are somewhat ambiguous. Firstly, the cut-off size of droplets for airborne transmission has always been controversial (8, 22, 23). The droplet nuclei, first defined by Wells (24), refers to the residues of droplets after complete evaporation. Centers for Disease Control and Prevention (CDC) defined the cut-off size of 5 µm for airborne transmission (25), and the threshold distance for airborne transmission is defined by the World Health Organization (WHO) as 1.0 m (26). However, it is known that droplet nuclei over 5 µm may also easily suspend and disperse over 1.0 m to cause transmission of respiratory disease, depending on the surrounding airflow conditions (27). Secondly, in some cases, airborne transmission was misinterpreted as merely long-range transmission (5), but the role of short-range airborne is quite important but usually ignored (28). Thirdly, close contact transmission (29, 30) can occur via multiple routes including short-range airborne (28), direct inhalation of droplets, deposition of droplets on facial membranes and secondary contact with droplets deposited on surfaces (18, 31).

Combining all these factors, we define the following transmission routes for our study (see Figure 1).

**Figure 1.**
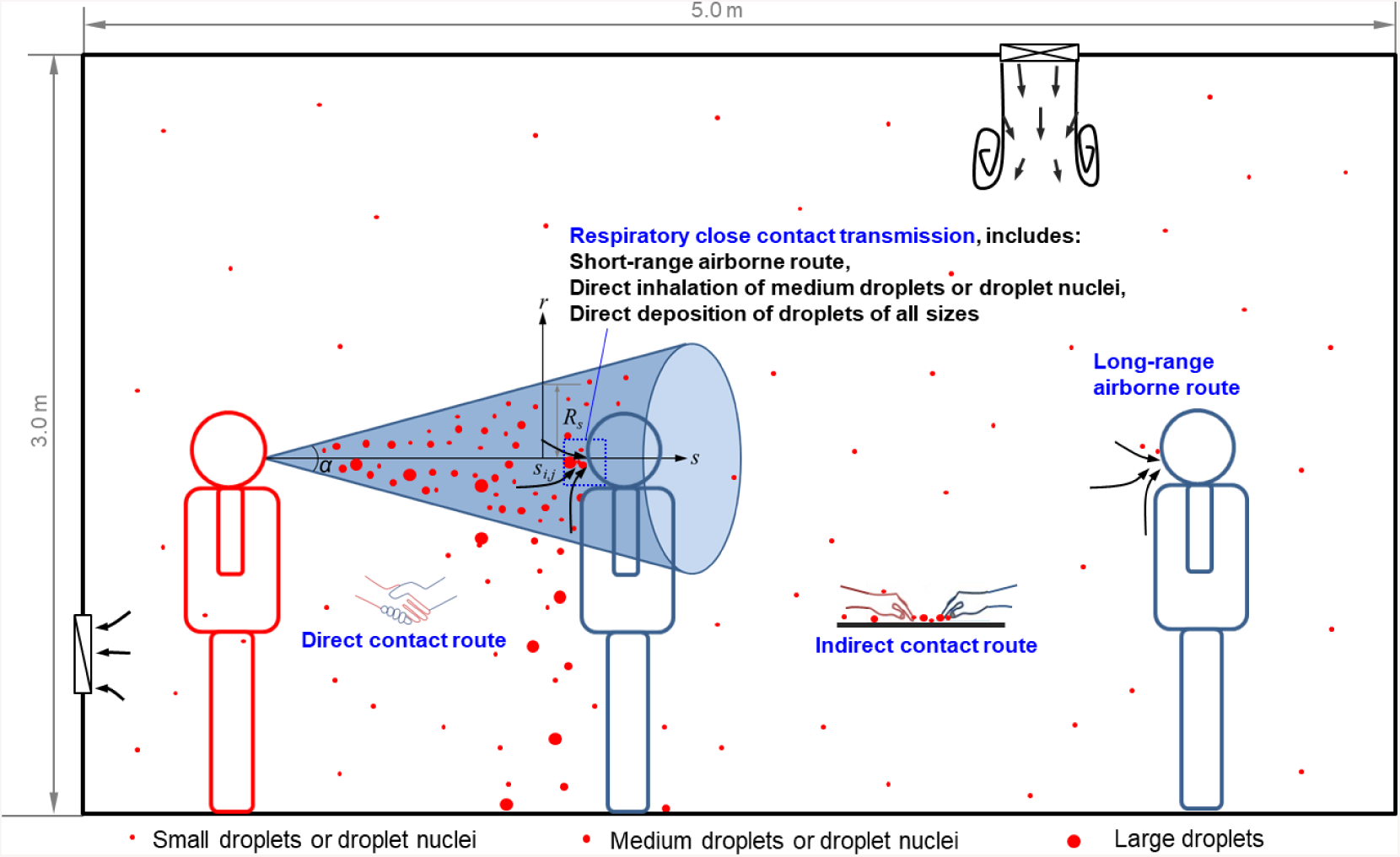
Illustration of different transmission routes of respiratory disease in indoor environments. *α* is the spreading angle of the idealized respiratory jet of the infector and *s*_*ij*_ is the exposure distance.

***Long-range airborne transmission*** – occurs when the susceptible shares the same indoor environment with the infector. For our model, infectious agents are assumed to be evenly distributed and reach a quasi-static equilibrium in the indoor environment. Detailed airflow patterns in individual environments are not considered.

***Respiratory close contact transmission* (*or direct spray route*)** - exposure of a susceptible individual within the exhalation jet of the infector. It includes three transmission mechanisms: (1) the *short-range airborne transmission*: exposure to droplets or droplet nuclei with a diameter less than a cut-off size of *d*_*a*_; (2) *direct inhalation of medium droplets or droplet nuclei* (*d*_*a*_ - 100µm in diameter); and (3) *direct deposition of droplets of all sizes*. Droplets larger than 100 µm cannot be inhaled but can cause infection by direct deposition on mucous membrane. Short-range airborne were considered distinct from direct inhalation, as larger droplets mostly deposit in the head airway and small droplets penetrate deeper in the lower respiratory airways, which may have different dose-response pattern. The relative facing orientation of the susceptible and the infector also affect exposure risk. In this study, we assume that respiratory close contact transmission only occurs when the breathing zone of the susceptible are inside the respiratory zone of the infector.

***Contact* (*or surface touch*) *transmission*** - is introduced by touching *contaminated surfaces* (*indirect contract route*) and *direct contact with the infector’s hand*. Surfaces are contaminated by direct deposition of respiratory droplets and/or by the hand of the infector. Here we only consider nonporous room horizontal surfaces and special hand contaminated surfaces (desks, door handles, etc.).

### Multi-route transmission model

Our multi-route model is developed with the capacity to be extended to evaluate infection risks in a location visiting network (32). We use *i* to denote a susceptible individual visiting location *k*, with infectors *j* = 1 … *N*. The total infection risk of individual *i* in location 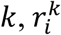, can be calculated combining the Wells-Riley equation (33) and the dose response model (34):

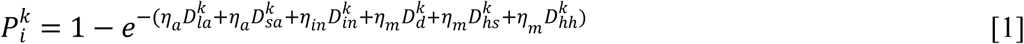

The effectiveness of the viral dose introduced by the various routes is potentially different (relates to the presence of virus receptors in different regions of the respiratory tract as well as facial membrane). Hence, we introduce three dose-response coefficients, namely *η*_*a*_, *η*_*in*_ and *η*_*m*_, to account for potential different dose-response rates via the airborne route, direct inhalation and exposure to facial membranes respectively. 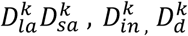 are exposure doses due to long-range airborne, short-range airborne, direct inhalation and direct deposition routes, respectively. 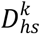 and 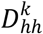 are exposure doses caused by hand contact with contaminated surface and direct hand to hand contact between the individual and the infector. The detailed mathematical derivation for each transmission route is provided in S1 Supplementary Material, and here we out outlined the final equations and important parameters.

***Exposure dose from long-range airborne route***, 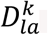, is calculated via estimating the steady-state concentration of droplet nuclei in room *k* and calculating the cumulative deposition infectious dose in susceptible’s respiratory tract. We use *d*_0_ to denote the original diameter of the droplet when it was generated from the mouth, and *d*_*r*_ to denote the diameter of droplet nuclei or residue (final size). Then the total exposure dose is a function of: droplet generation rate, *G*_*j*_(*d*_0_) from the infector; the respiratory deposition rate of droplet nuclei, *E*(*d*_*r*_); pulmonary ventilation rate, *p*; exposure time of the individual in the location 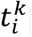; virus concentration in the respiratory droplet generated from the infector *j* on the day *T* of the course of infection *L*(*j, T*); room volume, *V*^*k*^, the air change rate (ACH) in the room, and particle loss rate due to the death in the air, *χ*_*a*_, and room deposition, *χ*_*d*_.

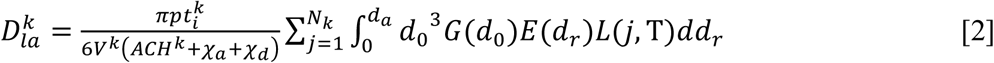

Although droplet nuclei (final) size, *d*_*r*_, is a function of its initial size, component, and relative humidity (35), the final droplet size is about the third of its original size after evaporation, *d*_*r*_ = 0.32*d*_0_, for a typical respiratory droplet (35, 36).

***Exposure dose from short-range airborne route*** *-* this risk is estimated differently due to its different transmission mechanism. We assume that breathing, talking, and coughing all generate a respiratory jet cone (see Figure 1) with a spreading angle of *α*. The concentration of droplet nuclei at distance *s*_*ij*_ can be estimated based on initial concentration and dilution rate along the cone. 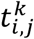 is the face-to-face contact exposure time of *i* and *j*. The concentration dilution factor at the distance *s*_*i,j*_ is a function of initial radius of the mouth open area, *R*_0_, spreading angle *α* and distance between two individuals.

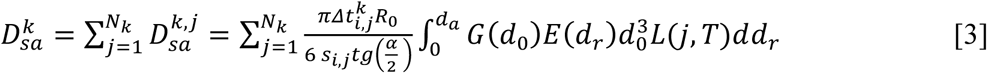

***Dose due to direct inhalation of medium droplets or droplet nuclei*** *-* 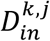. Medium sized droplets also travel in the respiratory jet, with larger ones falling out of the respiratory jet before reaching the breathing zone of the susceptible individual and smaller ones following the flow of the respiratory jet. The total infectious dose introduced by droplets with diameter between *d*_*a*_ and *d*_*b*,max_ (largest breathable droplet) can be calculated similarly as the short-range exposure of droplet nuclei. Similar to Equation [3], we have:

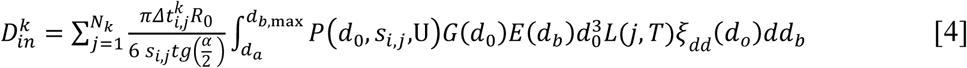

Different from Equation [3], Equation [4] has two additional parameters: *ξ*_*dd*_(*d*_*o*_)-virus concentration dilution factor in larger droplets; and *P*(*d*_0_, *s*_*i,j*_,U) - probability of droplets with an initial size of *d*_0_ to reach distance *s*_*i,j*_ before falling out of the respiratory jet at an initial speed (*U*). *ξ*_*dd*_(*d*_*o*_) is introduced as large droplets were often generated in the oral cavity (10), where antiviral substances are presented and viral load is often lower compared with smaller droplets generated from the lower respiratory tract, nasal cavity and pharynx. To simplify the modeling, we use a step function to represent *ξ*_*dd*_(*d*_*o*_)

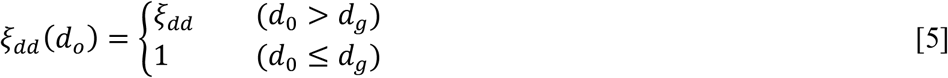

*P*(*d*_0_, *s*_*i,j*_, U) is affected by the mouth opening size, initial velocity of the jet, and the room temperature and humidity, which has been modeled by combing the buoyant round jet model and droplet evaporation and motion models (37).

***Dose due to direct deposition in the facial membranes*** - following the respiratory jet, both large and small droplets have the possibility to directly deposit on the susceptible’s facial membranes. This dose can be estimated as:

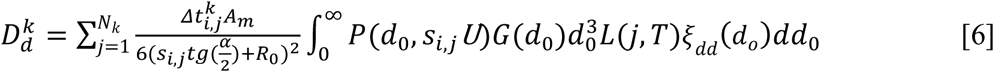

where *A*_*m*_is the area of facial membranes (surface area of the eyes, nostrils and lips).

***Exposure dose from hand-surface contact* (*touch*) *route*** *–* The virus on indoor surfaces is assumed to come from two sources: deposition of respiratory droplets and touch by contaminated hands of the infector. We assume that virus is uniformly distributed on hands, droplet and hand contaminated non-porous surfaces. Virus dose exposed to surfaces can be written as follows:

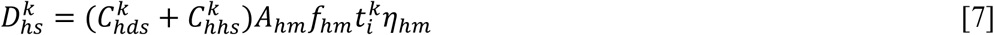

where, 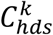 and 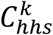 are the virus concentration in the hand of individual *i* by touching a droplet-contaminated and a hand-contaminated non-porous surface. *f*_*hm*_ is the frequency of the hand of touching facial membranes (eyes, nose and mouth). *η*_*hm*_ is the transmission rate of droplets from hand to facial membranes. *A*_*hm*_ is the contact area of hand to mucous membranes. By assuming that virus concentrations on those surfaces reach a steady-state, we have:

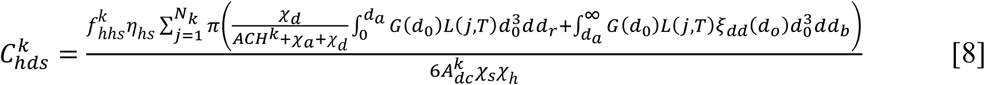

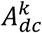 total droplet contaminated area in room *k*.

Virus dose introduced via touching a special hand-contaminated surfaces (i.e., door handles, desks), 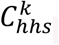, can be estimated via first estimate virus concentration on infector’s hand, and then virus concentration on the surface and finally virus concentration on the susceptible’s hand. This is expressed as a function of the virus death rate on the surface, *χ*_*s*_, and on hands, *χ*_*h*_; the frequency of hand touching the surface, 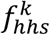 and facial membranes, *f*_*hm*_; virus transmission rate of between surface and hand per touch *η*_*hs*_ and the volume of contagious nasal discharge transported the infectors hands per touch of facial membrane *V*_*mh*_.

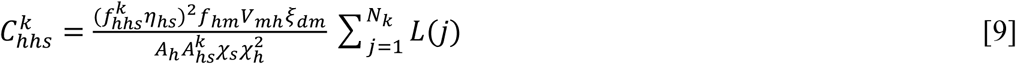

We used one additional parameter, *ξ*_*dm*_, to denote the virus concentration dilution rate in the nasal discharge compared with respiratory droplet. Covering mouth or nose while coughing was not considered as probability of covering and the dose of introduced virus are hard to estimate.

***Exposure dose from direct hand to hand route*** *–* Assuming the virus is uniformly distributed on hands. Virus dose exposed to hand can be written as follows:

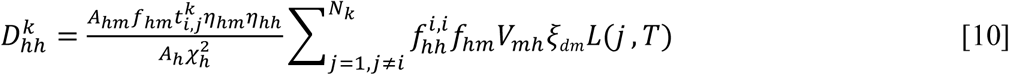

Here 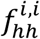 is the frequency of handshaking between *i* and the infector *j. η*_*hh*_is the virus transmission rate of a hand-hand touch.

### Modelling parameters and evidence sources

Room settings are illustrated in Figure 1. We consider a single room of 5(length)×5(width)×3(height) m^3^, with a ventilation rate in the range of 0.5-10 air changes per hour (ACH). One susceptible and one infector share the room for a 10-hour period with 1 hour face-to-face exposure when the susceptible is exposed directly in jet cone created by infector’s respiratory activities. Exposure distances were evaluated between 0.5-1.5 m of this close contact exposure. Virus load detected from samples of nose and throat swabs of influenza infectors is approximated according to 2009 Swine flu (38) (see S2 in Supplementary Material). Data of respiratory droplet generation rate and size distribution is estimated based on the study by Duguid (39) (see S3 in Supplementary Material). Large droplets viral dilution rate and cutoff size are largely unknown. In an unpublished data by Cowling and colleagues, only two positive samplers were found in saliva swabs from all 53 confirmed influenza A or B infectors. To be conservative, we assume that the virus load dilution rate, *ξ*_*dd*_ (*d*_*o*_), is 0.05 in respiratory droplets larger than *d*_*g*_. The travel distance and final size of respiratory droplets estimated using our droplet dispersion model are provided in S4 in Supplementary Material. Other parameters are listed in Table 1. Since droplet nuclei smaller than 5 µm in diameter are able to reach the pulmonary region (Figure S2) during inhalation and will suspend in the air for a long period (37), the threshold value of 5 µm diameter used as the cut-off size for airborne droplet *d*_*a*_. Sensitivity analysis were conducted with changing key impacting parameters. All modelling tasks were conducted using R version 3.6.1 (2019-07-05). Modelling code are provided in Supplementary R code.

**Table 1.** Input data for the multi-route transmission mode

| Parameter | Value | Source |
| --- | --- | --- |
| Room volume $V^k$, [m<sup>3</sup>] | 75 | Environment setting |
| Virus concentration in the droplet with an initial diameter $d_0$ and day T after symptomatic, $L(j, d_0)$ [TCID<sub>50</sub>/mL] | $10^{6.25}$ | (38), see S2 in Supplementary Material |
| Pulmonary ventilation rate, $p$ [m<sup>3</sup>/h] | 0.48 | (44) |
| Diameter of largest high viral-load, $d_g$ [μm] | 50 | Assumed (30-2000 included in sensitivity analysis) |
| Cut-off size for airborne droplets, $d_a$ [μm] | 5 | Assumed (5 for sensitivity analysis) |
| Initial speed of respiratory jet, $U$ [m/s] | 5 | (45, 46) |
| The dose effects of introducing infection through the airborne route, $\eta_a$ | 1 | (18, 19) |
| The dose effects of introducing infection through the large droplet direct inhalation, $\eta_{in}$ | 0.1 | Assumed (0.001-1 for sensitivity analysis) |
| The dose effects of introducing infection through the mucous membrane, $\eta_m$ | 0.01 | (15, 16) (0.001-1 for sensitivity analysis) |
| Virus concentration dilution rate in larger droplets $\xi_{dd}$ | 0.05 | Based on unpublished data (1 for sensitivity analysis) |
| Virus concentration dilution rate in nasal discharge $\xi_{dm}$ | 0.1 | Assumed (1 for sensitivity analysis) |
| Air change rate (ACH) | 1 | 10 L/s per person (0.5-10 for sensitivity analysis) |
| Virus death rate in the droplets nuclei at RH=24% and 7°C, $\chi_a$ [h<sup>-1</sup>] | 0.22 | (47) |
| Particle deposition lost-rate coefficient $\chi_d$ [h<sup>-1</sup>] | 2.2 | (48) |
| Virus death rate on hand, $\chi_h$ [h<sup>-1</sup>] | 12 | (49) |
| Virus death rate on non-porous surfaces, $\chi_s$ [h<sup>-1</sup>] | 0.18 | (49) |
| Frequency of hand of individual touching mouth and nose (the mucous membranes), $f_{hm}$ [h<sup>-1</sup>] | 5 | (18) |
| Frequency of individual touch a droplet contaminated non-porous surface, $f_{hds}^k$ [h<sup>-1</sup>] | 20 | Assumed |
| Frequency of individual touch a hand contaminated non-porous surface, $f_{hhs}^k$ [h<sup>-1</sup>] | 20 | Assumed |
| Frequency of shaking hand of $i$ and $j$, $f_{hh}^{i,j}$ [h<sup>-1</sup>] | 1 | Assumed |
| Transmission rate from hand to mucous membranes, $\eta_{hm}$ | 35% | Estimated from bacteria transmission (50) |
| Transmission rate between non-porous surfaces and hand, $\eta_{hs}$ | 8% | (49) |
| Virus transmission rate from hand to hand, $\eta_{hh}$ | 8% | Assumed |
| Spreading of the respiratory jet, $\alpha$ | 25° | (45, 46) |
| Radius of the mouth open area, $R_0$ [cm] | 1.0 | (45, 46) |
| Area of facial membranes of people (total surface area of the eyes, nostrils and lips), $A_m$ [cm<sup>2</sup>] | 10 | (18) |
| Hand area, $A_h$ [cm<sup>2</sup>] | 10 | (18) |
| Connection area of hand to mucous membranes, $A_{hm}$ [cm<sup>2</sup>] | 1 | Assumed |
| Large droplet contaminated area, $A_{dc}^k$ [m<sup>2</sup>] | 25 | Assumed |
| Area of special hand-contaminated surface in the room, $A_{hs}$ [m<sup>2</sup>] | 25 | Assumed |
| Volume of nasal discharge transported to hands of the infector per touch, $V_{mh}$ [mL] | 0.01 | Assumed |

## Results

Infection risk and contribution ratios by different transmission routes are evaluated under different modeling parameters. Figure 2 summarized results with the default modeling parameters, and five other scenarios when a certain transmission route dominates.

**Figure 2.**
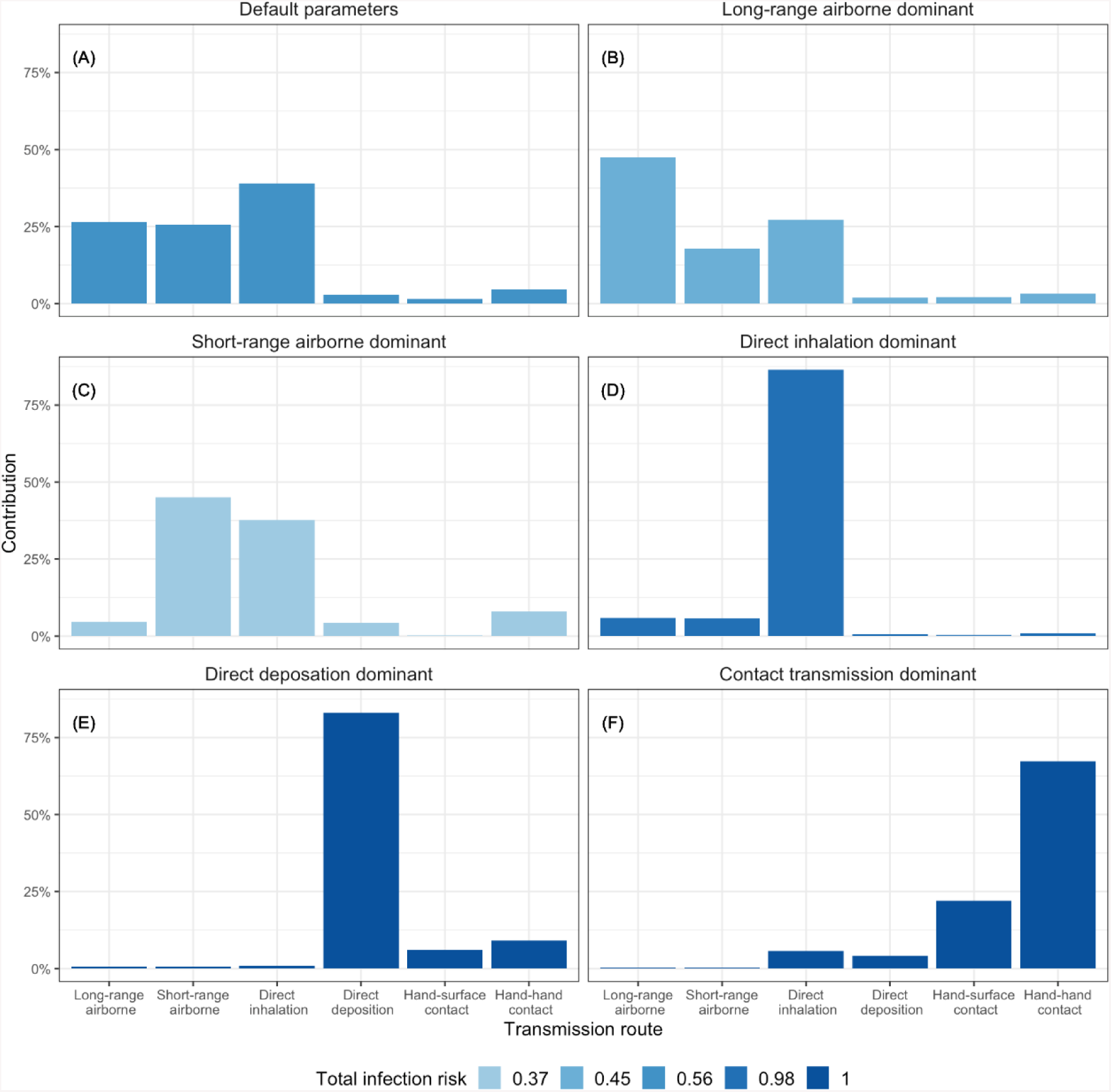
Total infection risk and relative contribution under different transmission route dominant scenarios. Note: parameter setting for the models are: (A) default, see Table 1; (B) face to face exposure time *t*_*i,j*_ = 0.5 h and ACH=0.5; (C) largest high viral-load droplet, *d*_*g*_ = 30 µm and room exposure time *t*_*k*_ = *t*_*i,j*_ = 1 (D) dose-response coefficient for direct inhalation, *η*_*in*_ = 1; (E) virus concentration dilution rate in larger droplets *ξ*_*dd*_ (*d*_*o*_) = 1 and the dose-response coefficient for membrane exposure *η*_*m*_ = 1; (F) all dose effects coefficient *η*_*a*_ = *η*_*in*_ = *η*_*m*_ = 1 and nasal discharge dilution factors *ξ*_*dm*_ = 1.

Overall infection risk is highest in the scenario displayed in Figure 2F, where all the dose-response coefficients are equal to 1 and the viral load in nasal discharge is the same as that in small respiratory droplets. This is only true when virus receptors are widely prevalent (in facial membrane, oral/nasal cavity upper/lower respiratory track) and nasal discharge and saliva are as contagious as small droplets; contact route dominates the transmission.

Figure 2E presents the scenario when there is no virus dilution in large droplets (*ξ*_*dd*_ (*d*_*o*_) = 1), and transmission is dominated by direct deposition. Virus concentration of different sized droplets remains unclear, although droplets smaller than 5µm in diameter are proved important virus carriers for influenza(40, 41). In scenario Figure 2D, when the dose-effect of direction inhalation is as high as airborne droplet inhalation (*η*_*a*_ = *η*_*in*_ = 1), transmission is dominated by direct inhalation; the total volume of medium sized droplets is larger although there are more smaller droplets (in number) generated.

When total exposure time in the room is the same as close-contact time and larger droplets (*d*_*g*_ > 30 µm) do not contain high concentration of virus, the dominating transmission route is short-range airborne transmission (Figure 2C). When the face-to-face exposure time is shorter (*t*_*i,j*_ = 0.5 h) and room ventilation is poor (ACH=0.5), long-range airborne transmission dominates.

We also evaluated the role of *da* (cut-off size for airborne particle) and *dg* (largest high viral-load droplet size) in impacting total infection risk (see Table 2). Total infection risk is higher with larger airborne cut-off size, particularly when the exposure distance is longer.

**Table 2.** Overall infection risk during the 10-hour exposure with varying exposure distance (*s*_*i,j*_), the largest high viral-load droplet size (*d*_*g*_) and cut-off size for airborne route (*d*_*a*_).

| Cut-off size for<br>airborne route | Largest high viral-load droplet size ($d_g$) | | | | | Largest<br>droplet<br>size |
| --- | --- | --- | --- | --- | --- | --- |
|  | 30 | 50 | 100 | 200 | 500 |  |
| Exposure distance $s_{i,j}=0.5$ m | | | | | | |
| $d_a=5\mu\text{m}$ | 0.62 | 0.71 | 0.74 | 0.77 | 0.97 | 1.00 |
| $d_a=10\mu\text{m}$ | 0.94 | 0.95 | 0.96 | 0.96 | 0.99 | 1.00 |
| Exposure distance $s_{i,j}=1.0$ m | | | | | | |
| $d_a=5\mu\text{m}$ | 0.49 | 0.56 | 0.58 | 0.58 | 0.61 | 0.70 |
| $d_a=10\mu\text{m}$ | 0.92 | 0.94 | 0.94 | 0.94 | 0.94 | 0.95 |
| Exposure distance $s_{i,j}=1.5$ m | | | | | | |
| $d_a=5\mu\text{m}$ | 0.46 | 0.52 | 0.54 | 0.54 | 0.54 | 0.54 |
| $d_a=10\mu\text{m}$ | 0.92 | 0.93 | 0.93 | 0.93 | 0.93 | 0.93 |

Note: other parameter settings were listed in Table 1

Another key impact factor is the largest high viral-load droplet size *dg*. As *dg* becomes larger, total infection risk increases dramatically when exposed at a closer distance (e.g., when the exposure distance is 0.5m, infection risk changed from less than 0.5m to 1.0m when *dg* increased from 30 to largest droplet size). This is due to higher contribution from direct droplet inhalation and large droplet deposition (see Figure 3). Increasing exposure distance also reduces total risk by reducing direct droplet inhalation and deposition (Table 2 and Figure 3)

**Figure 3.**
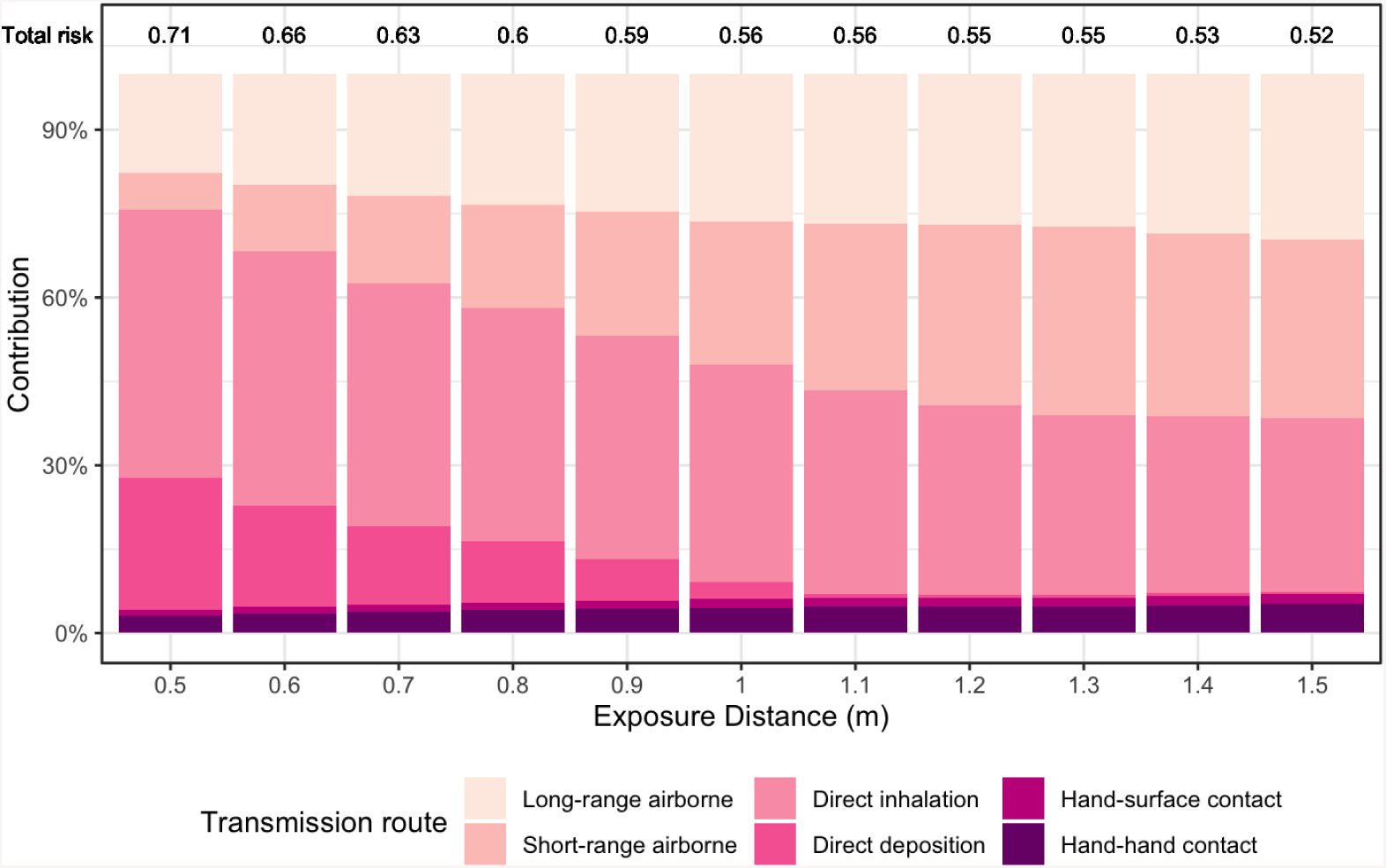
Effect of close-contact exposure distance on contribution ratios of different transmission routes. Note: parameter setting see Table 1.

Effect of ventilation rates on contribution ratios of the long-range airborne route is demonstrated in Figure 4. In the long-range airborne transmission dominant scenario (face-to-face exposure time *t*_*i,j*_ = 0.5), with the increase of the air change rate from 0.25 (18.75 m3/h) to 10 ACH, the total infection risk decreases by ~40% (85% reduction in infection risk from long range airborne route).

**Figure 4.**
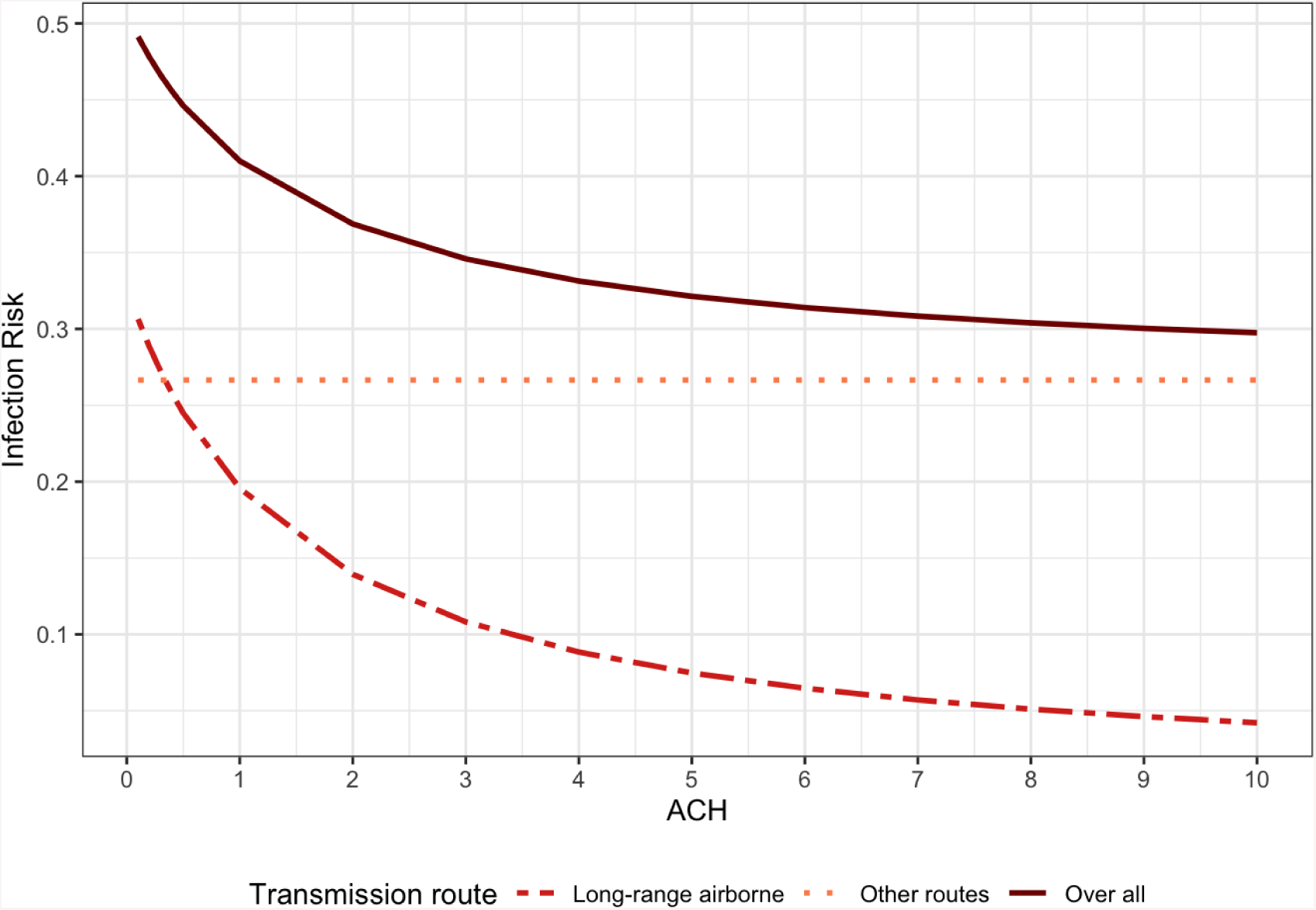
Effect of ventilation rates on contribution ratios of different transmission routes. Note: face to face exposure time 30 minutes and other parameter settings see Table 1.

We also conducted sensitivity analyses with variable dose-effect coefficients. When the dose effect of mucous membrane transmission was assumed larger, direct hand-to-hand contact transmission became more important, particularly when the virus concentration is also high in nasal discharge (see Figure 5). Similarly, when the dose-effect coefficient of direct inhalation route is high, the infection risk is dominated by direct inhalation particularly when viral load is also high in larger droplets (see Figure 6)

**Figure 5.**
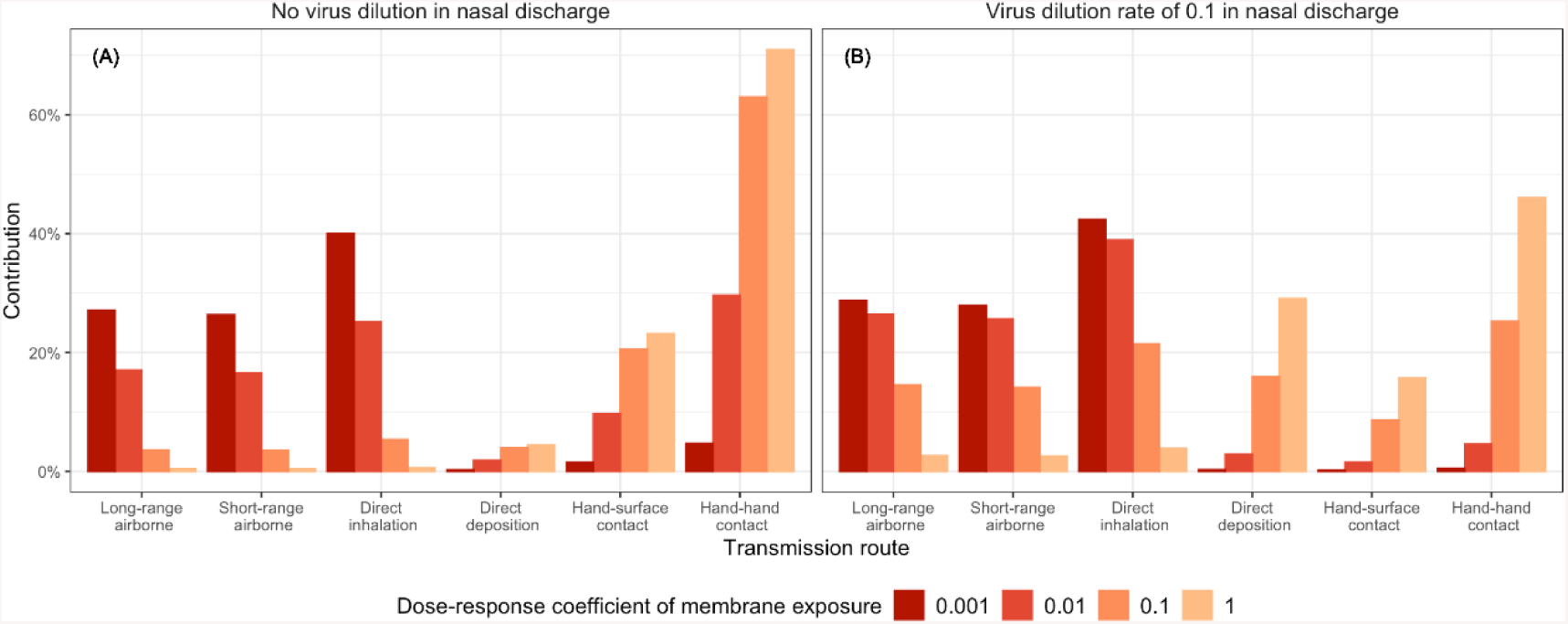
Contribution ratios of different transmission routes with varying dose-response coefficient for membrane exposure, *η*_*m*_, and nasal discharge virus dilution rate, *ξ*_*dm*_. Other parameters were set the same as listed in Table 1.

**Figure 6.**
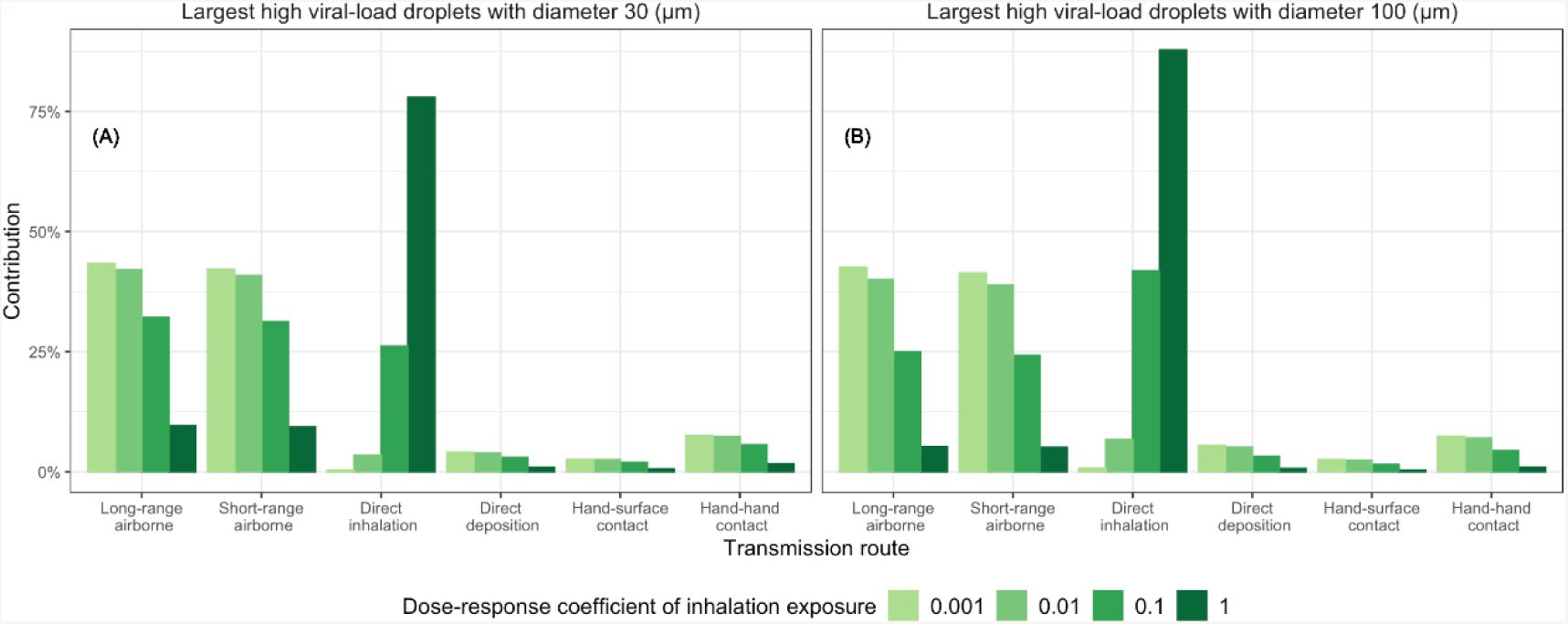
Contribution ratios of different transmission routes with varying dose-response coefficient for inhalation exposure, *η*_*in*_, and diameter of largest high viral-load droplet, *d*_*g*_. Other parameters were set the same as listed in Table 1.

## Discussion

Given the recent coronavirus pandemic, this study provided a timely and novel approach in evaluation transmission mechanisms of respiratory infection. We first proposed a new definition of multi-route transmitted respiratory infection and developed a full mathematical model to evaluate contributions of each transmission route. We illustrated that each transmission route can dominate the total infection risk under different virological, environmental and behavior settings. Importantly our findings dissipate the traditional dichotomy of respiratory infection being transmitted by either close-contact or airborne routes. All these factors should be taken into consideration conjunctively to design intervention methods.

Our model may also explain the inconsistencies of research on the role of airborne transmission in historical outbreaks (8, 11, 12). The airborne transmission can be dominant when the ventilation was poor. For example, in the 1977 Alaska aircraft influenza outbreak all passengers were exposed to a constant coughing patient an enclosed aircraft without ventilation for more than 3 hours in the (11). However, when the ventilation rate is high, or room exposure time is short, the contribution of airborne transmission can be insignificant, such as in the influenza tour group outbreak in 2009 in China (12).

Compared with the models previously developed (18, 19), our model provides substantial advancement in modelling the complex droplet evaporation, transportation and deposition process, which equipped us with the ability to distinguish between long-range and short-range airborne, direct inhalation and direct deposition transmission routes. Unlike other studies, we have also evaluated how the key parameters, such as dose-response coefficient of different routes, the exposure distance, room ventilation rate and viral-load in large droplets and nasal discharge, impact the total infection risk and the relative contribution from each route. This also shed the light on important factors to consider when evaluating transmission of a novel respiratory infection, such as prevalence of virus receptors in different parts of respiratory track and difference of viral load in lung fluid, sputum, nasal discharge, saliva etc.

Another important difference in our model compared to previous work is that we considered the evaporation process of respiratory droplet as well as the *airborne* ability related to the final size rather than initial size. Respiratory activities mostly generate substantial number of droplets between 0 and 50 µm. Droplets with the initial diameter of 10 µm (with a volume 1000 times larger than a 1 µm droplet) will evaporate quickly to a diameter of about 3 µm, which can then penetrate into the lower respiratory track. Using an airborne cut-off size of *d*_*a*_=5 µm, about 60% of respiratory droplets generated (*d*_0_=15 µm) can be evaporated and suspended in the air and introduce long-and short-range airborne infection, which is largely underestimated in other studies.

Cut-off size is also critical factor. WHO (26) employed 5 μm to divide airborne and large droplets, while Nicas et al. (42) adopted 10μm. The simulations by Xie et al. (43) and Wei and Li. (37) suggested that droplets with an initial diameter up to 60 μm could all disperse in a patient ward. In reality, the *airborne* ability of droplets or droplet nuclei may not be a simple cut-off value, and it depends on many other factors such as the background room air speed and its turbulence, thermal stratification and air distribution. Further studies are needed to model these factors to account for the long-range airborne transmission rate.

Although we have differentiated possible dose-response difference between airborne droplets and larger inhalable droplets, we did not model their deposition coefficient in different regions of respiratory tract, which can be a key impacting factor in total transmission. If the virus receptor only presented in the pulmonary region, all transmission routes except the airborne route have been overestimated as only particles less than 10 μm can penetrate the larger respiratory airways. In our model, breathing was also not included in the droplet generation process, and initial respiratory jet velocity differences between respiratory activities were not considered. Further development is needed to allow the model to separately consider different types of respiratory activities including breathing, talking and coughing.

We have used parameters such as viral load and virus survival rate for influenza; however, this model can be effectively adapted to model all multi-routes transmitted respiratory diseases such as SARS-CoV-2. It can also be used to evaluate intervention methods such as increasing exposure distance and wearing masks.

This study may provide theoretical guidance for future research that explores transmission mechanism and inform policy makers for efficient infection control. To this end, we have made all the simulation codes freely available in an open access code-repository.

## Conclusions

We have developed a multi-route mathematical model to distinguish contributions of each route in influenza transmission under different exposure settings. It is highlighted that the transmission mechanism is complicated and all different transmission routes may dominate the total infection risks. Therefore, recommendations of individual level interventions for influenza and other related respiratory illnesses should be guided by all virological, behavioural and environmental factors.

## Data Availability

The online code of the multi-route transmission model is available at GitHub.

https://carolinexgao.github.io/Multi-route/Simulation_submission.html

## Acknowledgement

This work was financially supported by the National Natural Science Foundation of China (51278440) and the Research Grants Council of Hong Kong’s Collaborative Research Fund (C7025-16G), the Fundamental Research Funds for the Central Universities, and the HKU-Zhejiang Institute for Research and Innovation (HKU-ZIRI) Seed Funding Programme.

## Author contributions

C.X., Y.L., J.W. and B.J.C conceived the study and wrote the paper which was reviewed by all authors. C.X.and J.W. performed the modelling, and S.C., M.H. and L.W. assisted in parameterization and data analysis.

## Additional information

The online code of the multi-route transmission model is available at GitHub: https://carolinexgao.github.io/Multi-route/Simulation_submission.html

